# Work-Related Injuries in the Australian Education Sector: A Retrospective Cohort Study

**DOI:** 10.1101/2022.09.10.22279808

**Authors:** Fatimah M Al Afreed, Tyler J Lane, Shannon E Gray

## Abstract

**Introduction:** Educators are exposed to several work-related hazards. Evidence suggests musculoskeletal pain, psychological distress, and student-inflicted violence-related injuries are common. However, there is little evidence on the burden of workplace injury among Australian educators.

**Aim:** To compare incidence of injury claims and duration of compensated time off work between educators and non-educators, and associated factors.

**Methods:** Retrospective cohort study of 1,559,676 Australian workers’ compensation claims, including 84,915 educator claims, lodged between July 2009 to June 2015, from the National Dataset for Compensation-based Statistics. Cases were included if aged 18+ years and working in the education sector less than 100 hours per week. Negative binomial regression models estimated the relative risk of making a compensation claim and survival analyses calculated disability duration within educators by sex, age, injury type and mechanism, socioeconomic area, remoteness, and jurisdiction.

**Results:** Compared to non-educators, educators had lower rates of injury claims and shorter disability durations. However, educators had a higher rate of claims for mental health conditions and assault, with the highest risk being among those in special education and education aides. Among educators, injury claim rates were highest among special educators, education aides, and secondary educators.

**Discussion and Conclusion:** Though surveys indicate Australians in the education sector have higher incidences of work-related injuries, this study found lower incidence of injury claims and shorter disability durations than others. Educators’ injury-reporting and absenteeism behaviors may be constrained by ethical, social, and administrative attitudes. Educators had higher rates of claims for mental health and assault-related injury, particularly special educators, and education aides, which suggests a need for targeted prevention efforts.

## Introduction

Education sector workers are exposed to various hazards that put them at risk for physical and psychological injury [1], including long periods of standing and sitting, lifting heavy loads, high workload, and violence in the workplace [2,3]. Evidence suggests that injuries are common among educators, particularly musculoskeletal disorders [4]. Globally, between 39% and 95% of teachers reported musculoskeletal disorders [5–7] and two-thirds of secondary school teachers experienced disabling pain [8]. Low-back, neck, and upper-limb pain are the most commonly reported musculoskeletal disorders, which have been attributed to workplace demands like prolonged overhead writing [7], sitting, and standing [3]. Psychosocial factors, depression, weight, older age, being female, poor posture, high body mass index and longer teaching duration were significantly associated with reported musculoskeletal injury [2–4,6–12].

Educators face numerous psychosocial stressors including rapid curricular restructuring, increasing workload due to growing obligations and standardized testing, hostile workplace environment and bullying, and increased disorderly classroom behavior [13–16]. Continuous system changes, uncertainties due to low job security, high expectations, and environment also contribute to educators’ stress [17,18]. Stress is linked to dissatisfaction at work, depression, anxiety disorders, and psychosomatic manifestations, all of which have detrimental impacts on health and quality of work [17].

A survey among secondary schoolteachers in Hong Kong found a third experienced stress, a third had symptoms of depression, and over half showed symptoms of severe anxiety [19]. Several studies suggest that a substantial number of teachers continue working with chronic burnout due to lack of a suitable alternative employment [18,20,21]. An Australian study found a link between an ever-growing teachers’ individual workload and development of exhaustion and disengagement, the key dimensions of burnout [15]. While educators may have an elevated risk of developing mental health conditions [17–21], a review of the existing literature did not find evidence that educators report higher rates of mental illness compared to other occupation groups [22]. However, there is limited evidence within the Australian context [23].

Workplace violence can have severe physical and psychological implications on the wellbeing of educators and students alike. In Minnesota, student-inflicted injury accounted for 26% of educators’ injury claims and 8.6% of compensated disability duration [24]. Non-physical violence like verbal abuse, bullying, and sexual harassment had an annual prevalence of 38 in 100 educators, more than four times that of physical assaults (8.3 per 100) [25]. The risk has been found substantially higher among special education teachers [24–26] and teaching assistants [24]. In Australia, between 68% and 85% of surveyed Australian primary and secondary educators reported experiencing violence (bullying and harassment) inflicted-by students and parents [27,28].

While the current literature suggests a link between working in education and increased rates of physical and psychological injury, most originate from countries that are systematically different from Australia. The apparent increase of work-related injuries among educators and lack of comparable evidence indicate that an appraisal of the Australian educators’ experience is necessary. This paper aims to answer the following questions within the Australian context:

1. Do educators have higher risk of occupational injury than non-educators?
2. Do injured educators take more time off work than other sectors?
3. What factors are associated with occupational injuries and time off work among educators?

## Methods

### Setting

Around 94% of the Australian workforce is covered by compulsory workers’ compensation [29], a government-mandated and regulated insurance system that provides benefits to workers after work-related injury or illness. There are 11 major workers’ compensation schemes across Australia: one in each of the six states and two territories, and three Commonwealth systems: Comcare for federal government employees that also issues self-insurance licenses to large, national self-insured businesses, Seacare for seafarers, and a Department of Veterans’ Affairs system for military veterans; of these, only Comcare is included in this study.

### Data

This study utilizes the National Dataset for Compensation-based Statistics (NDS), a dataset compiled by Safe Work Australia from jurisdictional-level administrative workers’ compensation claims data [30]. The dataset includes information about the worker (e.g., age, sex), injury (e.g., nature, location), employer (e.g., industry size) and claim (e.g., duration). The latest available dataset was updated to June 2017. Safe Work Australia also supplied denominator data of estimates of covered workers and number of hours worked by financial year, age group, sex, industry, and occupation [31].

The study includes data from claims lodged between July 2009 to June 2015 to allow at least two years of follow-up for time loss calculations. Educators were identified based on the Australian and New Zealand Standard Classification of Occupation (ANZSCO) [32], non-educators included all other occupations; which enabled a realistic measure of the impact of working in the education sector on work-related injury reports compared to all other occupations. Cases were excluded if a worker was aged <18 years or had abnormal and unrealistic working hours (≥100 hours).

#### Dependent variables

The primary outcomes are injury rate per 1,000 covered workers and disability duration measured as the cumulative number of weeks compensated (hours compensated divided by pre-injury weekly hours worked). While this underestimates actual time off work, it is the most accurate indicator when using administrative data [33,34]. Disability duration was right-censored at 104 weeks to standardize follow-up.

#### Independent variables

Independent variables included factors previously associated with duration of time loss including age, sex, occupation, nature of injury, bodily location, socioeconomic status (SES), remoteness, jurisdiction, and whether the employer paid for system coverage (scheme insured) or managed their own claims (self-insured) [34,35].

Occupations were grouped into educators and non-educators. Educators included: childcare and early learning, primary, secondary, tertiary, vocational education, special education, education aides, and principals based on a modification of the *Australia and New Zealand Standard Classification of Occupations* (ANZSCO) [32] detailed in Appendix 1. Non-educators were comprised of all other occupations.

Age at the time of injury was categorized (18-24, 25-34, 35-44, 45-54, 55+ years). Injury groups were based on a modified version of the Type of Occurrence Classification System (TOOCS) version 3.1 [36]. Musculoskeletal conditions were categorized by bodily location (upper, lower, other).

Index of Relative Socio-economic Advantage and Disadvantage (IRSAD) was used to categorize residential postcodes into top quintile (“Most Advantaged”), bottom quintile (“Most Disadvantaged”), and the middle three quintiles (“Middle 3 Quintiles”) as an indicator of SES [37]. Remoteness was determined by Accessibility/Remoteness Index of Australia (ARIA) [38] which ranks residential postcodes by remoteness (five-point scale from “major cities” to “very remote”). Jurisdiction was categorized based on the scheme under which the injured worker was compensated.

### Analysis

Preliminary analysis revealed postcode-linked indicators (SES and remoteness) values were missing for 7.3% of all cases and 14% of educators. Missing values were strongly predicted by jurisdiction. Multiple imputations were applied based on the total proportion of cases with missing values (rounded up to 8 imputations). More than 80% of cases from the Australian Capital Territory’s private scheme were missing postcodes, which was therefore excluded.

Analyses focused on two main outcomes: injury claim rate and disability duration, relative to all other occupations. Descriptive statistics including number and proportion of claims, age group, sex, nature of injury, SES, remoteness, and jurisdiction, along with mean disability duration (and standard deviation) for educators and non-educators, and for educator subgroups. Rate of claims were compared using a negative binomial regression with a logged denominator offset. Rather than adjusting for condition type, each condition type was evaluated in sub-analyses, e.g., analysis focused on the rate of claims for musculoskeletal conditions, mental health, assault injuries. SES and remoteness variables were excluded from this analysis as denominators could not be calculated. Outputs are reported as incident rate ratios (IRR) with corresponding 95% confidence intervals.

Disability durations were compared using Cox proportional hazards survival analyses. Outputs are reported as hazard ratios (HR) with corresponding 95% confidence intervals. Lower HRs indicate reduced probability of exiting the compensation system at any point, a proxy for longer disability duration [34]. Both negative binomial and Cox regressions were repeated on only educators to determine which educator subgroups and independent variables had the highest claim rates and longest disability durations.

Analyses were conducted in R using R Studio.

### Ethical Approval

This study received ethics approval from the Monash University Human Research Ethics Committee (MUHREC) (CF14/2995 – 2014001663).

## Results

### Educators’ characteristics

Between July 2009 to June 2015, 84,915 claims were made by educators, comprising 5.4% of total claims (Table 1). Almost four-fifths of educator claims were from females, compared to a third of non-educator claims. Twenty-nine percent of educator claims were from secondary education, 22.3% primary education and 17.0% education aides.

**Table 1:**
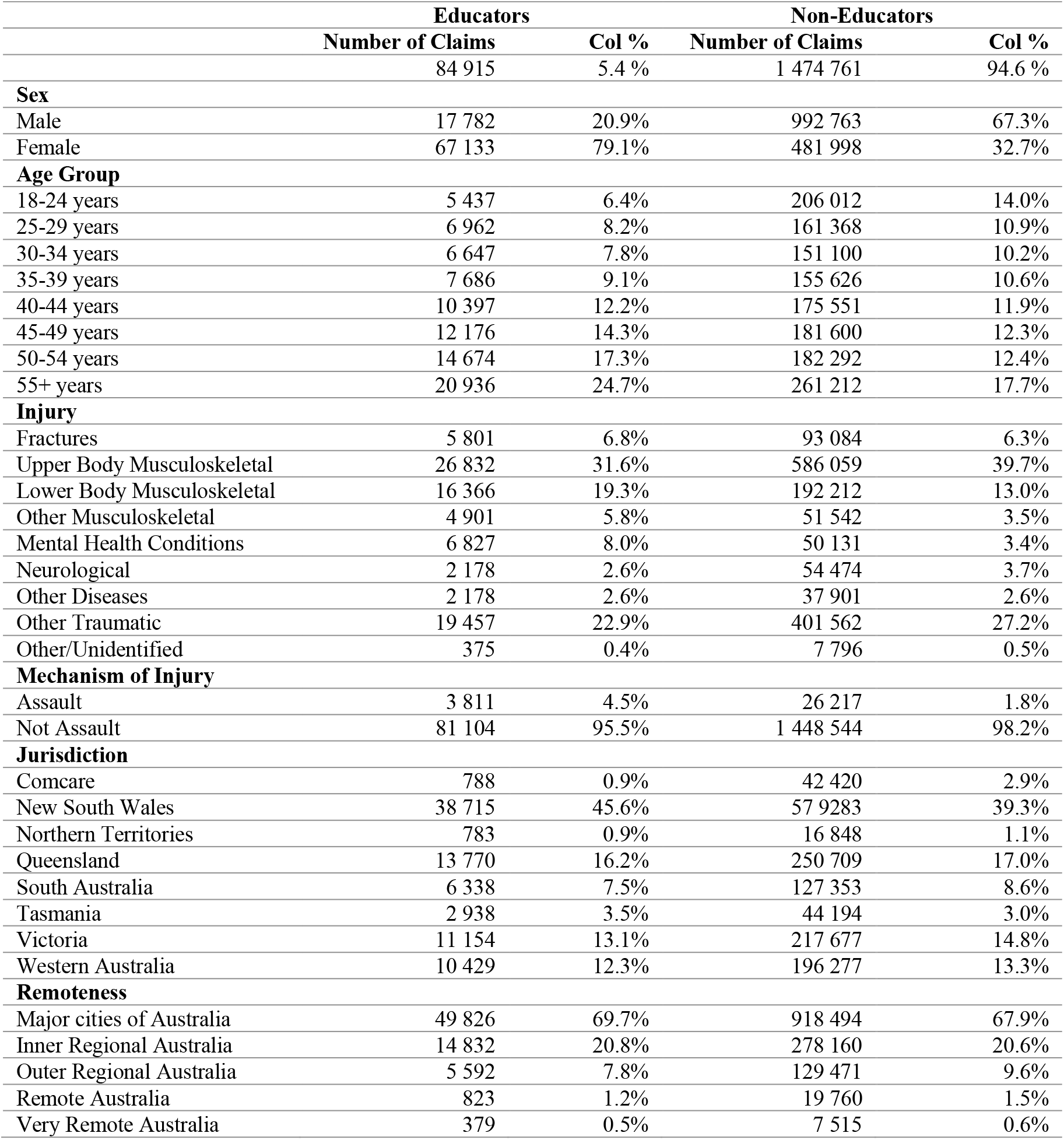
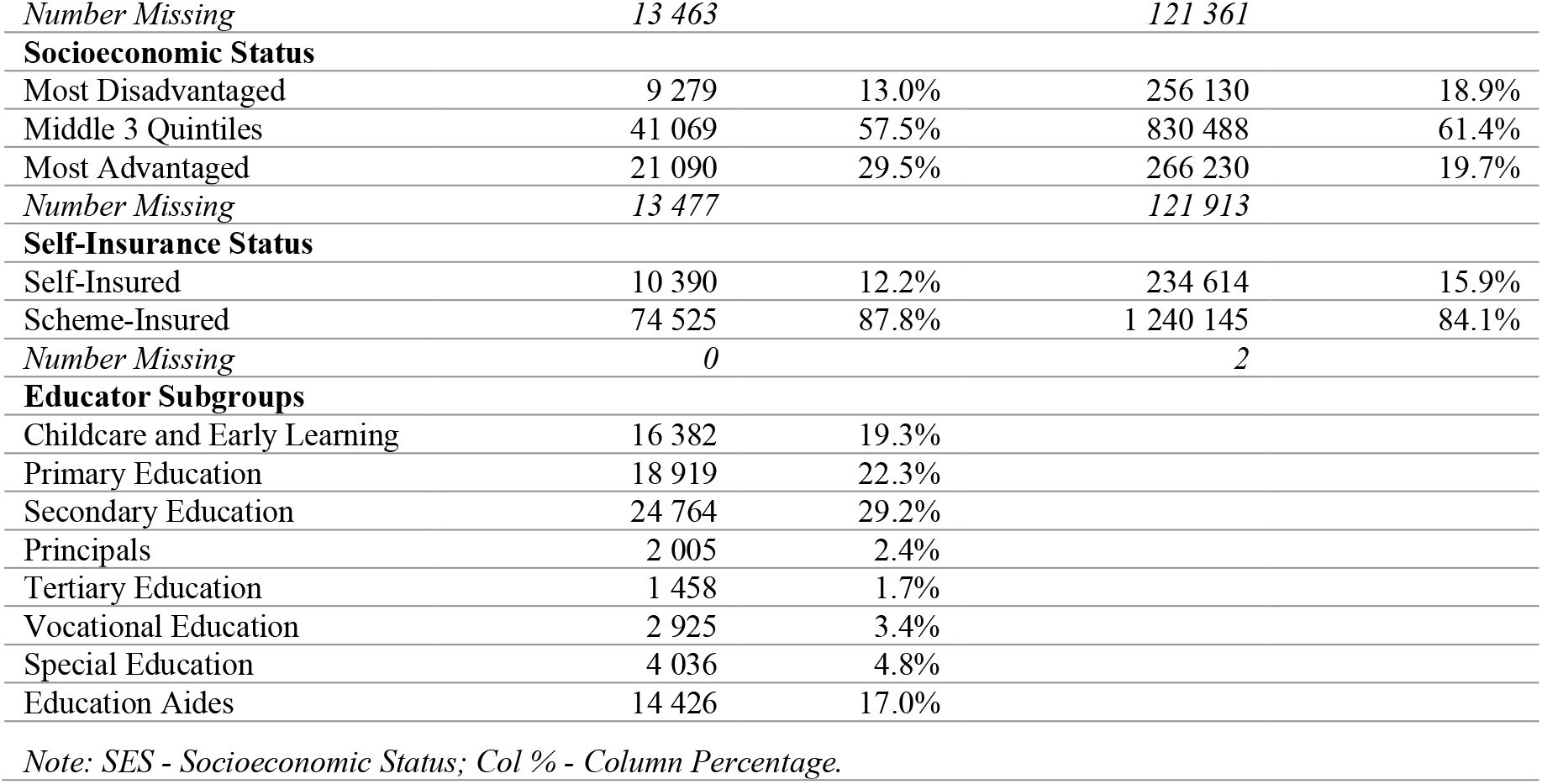
Frequency and proportion of claims among educators and non-educators by age group, sex, injury, jurisdiction, remoteness, SES, self-insurance status, and educator subgroups.

Claims for upper body musculoskeletal injuries were most common among educators (31.6%), though this was a lower proportion than in non-educators (39.7%). Lower body musculoskeletal injuries were also common (19.3% of educators’ claims and 13.0% of non-educators’ claims). Assault-related claims accounted for 4.5% of educator claims, but less than 2% of non-educators.

New South Wales had the highest proportion of claims for both educators (45.6%) and non-educators (39.3%). About 70% of both educators’ and non-educators’ claims were reported in major cities, and more than half of all injured workers were within the middle 3 quintiles of socioeconomic advantage.

Educators had shorter disability durations than non-educators, with mean time loss 6.8 weeks (standard deviation 23.0 weeks) for educators, and 10.7 weeks (standard deviation 33.6 weeks) for non-educators.

### Injury claim rates

The injury claim rate was 41% lower among educators than non-educators (Incidence Rate Ratio [IRR]: 0.59; 95% Confidence Interval [CI]: 0.54-0.66) (Table 2). However, educators had a 33% higher rate of mental health claims (IRR: 1.33; 95%CI: 1.18-1.49), and 74% higher rate of injury claims due to assault (IRR: 1.74; 95% CI: 1.55-1.94). Educators had a significantly lower claim rate for all other conditions except lower body musculoskeletal, which had no detectable difference (p = 0.372).

**Table 2:**
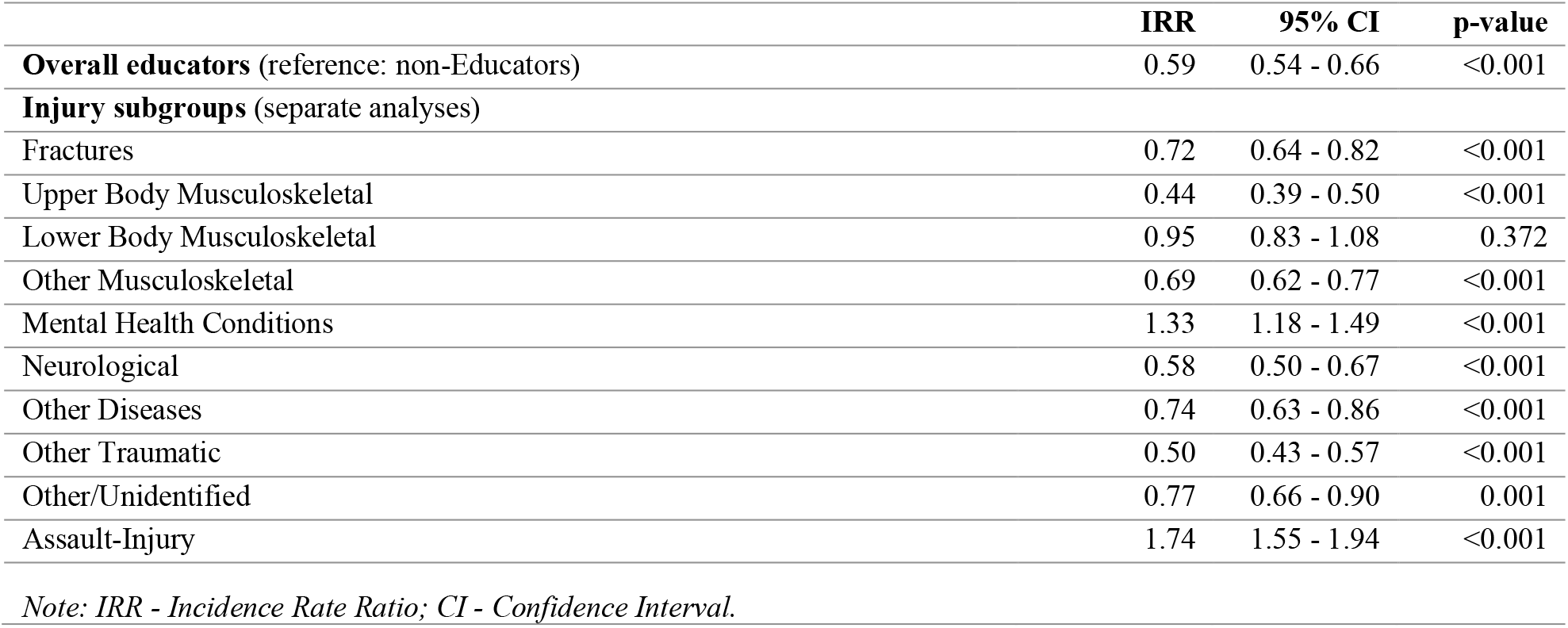
Negative binomial regression results comparing injury claim rate between educators and non-educators.

Among educator subgroups, the highest rates of occupational injury claims were in special education (IRR: 2.46; 95% CI: 2.19-2.77) and education aides (IRR: 1.58; 95% CI: 1.41-1.76) and lowest in tertiary education (IRR: 0.19; 95% CI: 0.16-0.21). While all jurisdictions had lower rates than Comcare, Victoria had the lowest rate (IRR: 0.35; 95% CI: 0.30 – 0.40) These results are summarized in Table 3.

**Table 3:** Negative binomial regression results for factors predicting overall claim rates among educators.

|  |  |  |  |
| --- | --- | --- | --- |
|  | IRR | 95% CI | p-value |
| **Educator Subgroup** (reference: Secondary Education) |  |  |  |
| Childcare and Early Learning | 1.13 | 1.01 - 1.26 | 0.037 |
| Primary Education | 0.64 | 0.57 - 0.71 | <0.001 |
| Principals | 0.55 | 0.48 - 0.62 | <0.001 |
| Tertiary Education | 0.19 | 0.16 - 0.21 | <0.001 |
| Vocational Education | 0.49 | 0.44 - 0.56 | <0.001 |
| Special Education | 2.46 | 2.19 - 2.77 | <0.001 |
| Education Aides | 1.58 | 1.41 - 1.76 | <0.001 |
| **Sex** (reference: Female) |  |  |  |
| Male | 1.00 | 0.94 - 1.07 | 0.887 |
| **Age group** (reference: 18-24 years) |  |  |  |
| 25-29 years | 1.31 | 1.14 - 1.51 | <0.001 |
| 30-34 years | 1.50 | 1.31 - 1.72 | <0.001 |
| 35-39 years | 1.58 | 1.38 - 1.81 | <0.001 |
| 40-44 years | 1.92 | 1.68 - 2.20 | <0.001 |
| 45-49 years | 2.30 | 2.01 - 2.63 | <0.001 |
| 50- 54 years | 2.42 | 2.12 - 2.76 | <0.001 |
| 55+ years | 2.42 | 2.12 - 2.76 | <0.001 |
| **Jurisdiction** (reference: Comcare) |  |  |  |
| New South Wales | 0.90 | 0.78 - 1.03 | 0.113 |
| Northern Territories | 0.44 | 0.37 - 0.51 | <0.001 |
| Queensland | 0.45 | 0.39 - 0.52 | <0.001 |
| South Australia | 0.65 | 0.56 - 0.76 | <0.001 |
| Tasmania | 0.90 | 0.78 - 1.05 | 0.174 |
| Victoria | 0.35 | 0.30 - 0.40 | <0.001 |
| Western Australia | 0.67 | 0.58 - 0.78 | <0.001 |

*Note: IRR - Incidence Rate Ratio; CI - Confidence Interval.*

#### Rate of claims for assault and mental health conditions by occupation group

Further analysis was conducted to identify factors associated with increased rate of mental health conditions and assault injury claims as shown in Table 4. Male educators had a 14% lower rate of injury claims for assault (IRR: 0.86; 95% CI: 0.75-0.99). The mental health claim rate was highest in the 50-54 years age group (IRR: 5.30; 95% CI: 4.18-6.74), and highest assault risk was in 45-49 years group (IRR: 1.67; 95% CI: 1.29-2.18). Special educators had the highest rate of injury claims for assault (IRR: 8.46; 95% CI: 6.93-10.31) and mental health (IRR: 1.17; 95% CI: 0.98-1.40), though the latter was non-significant compared to the reference category, secondary education. Education aides also had higher rates of injury claims for assault (IRR: 3.68; 95% CI: 3.05-4.43). Other educator subgroups generally had significantly less risk than secondary educators, indicating that the secondary education group has an increased risk compared to other educator subgroups. Tertiary educators had the lowest rate of injury claims for mental health conditions (IRR: 0.14; 95% CI: 0.11-0.17) and assault (IRR: 0.07; 95% CI: 0.03-0.13). All jurisdictions had lower claim rates than Comcare, Queensland had the lowest rate of mental health claims (IRR: 0.26; 95% CI: 0.20-0.33), and New South Wales had the lowest assault claims rate (IRR: 0.21; 95% CI: 0.15 – 0.29).

**Table 4:**
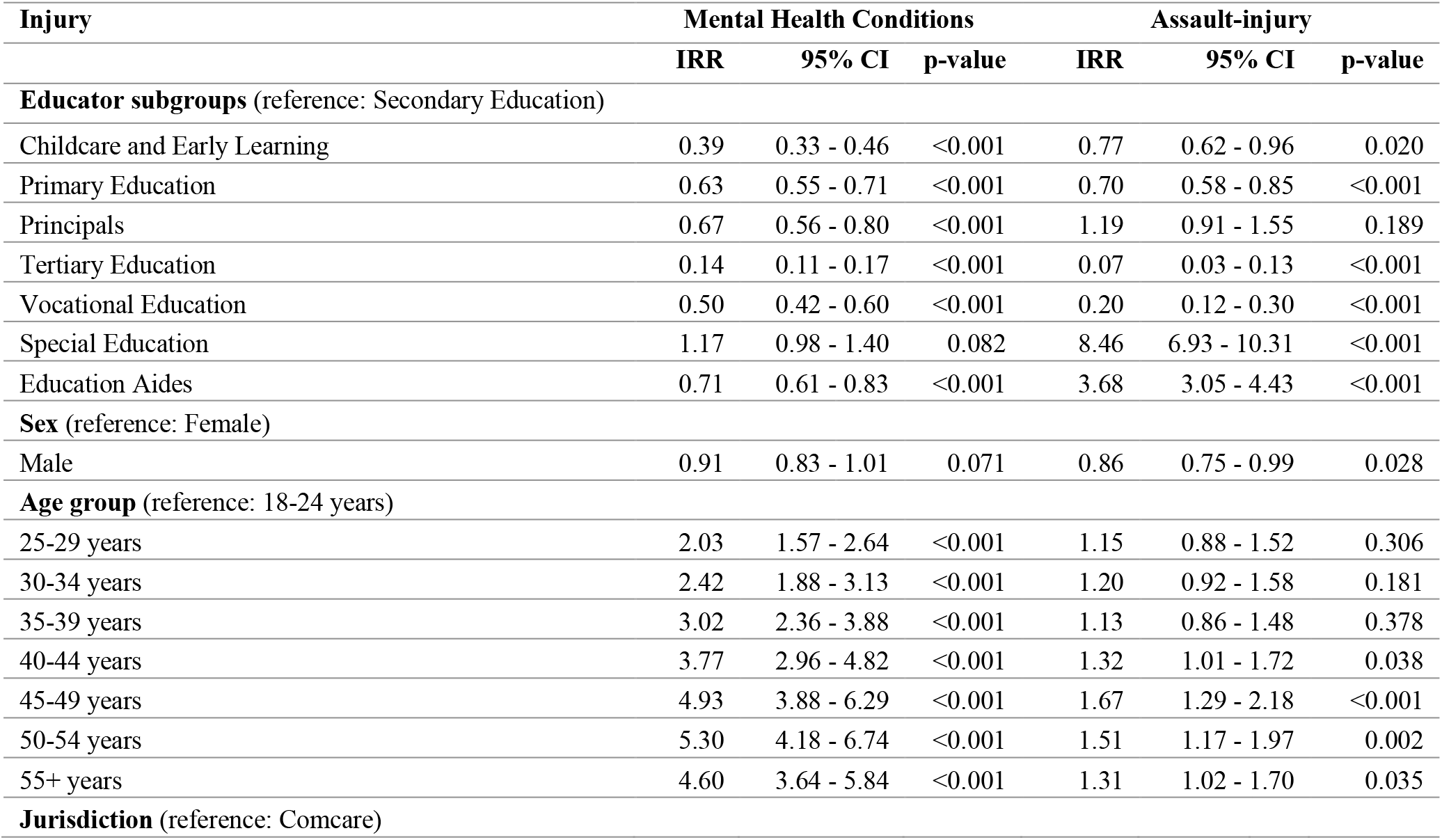
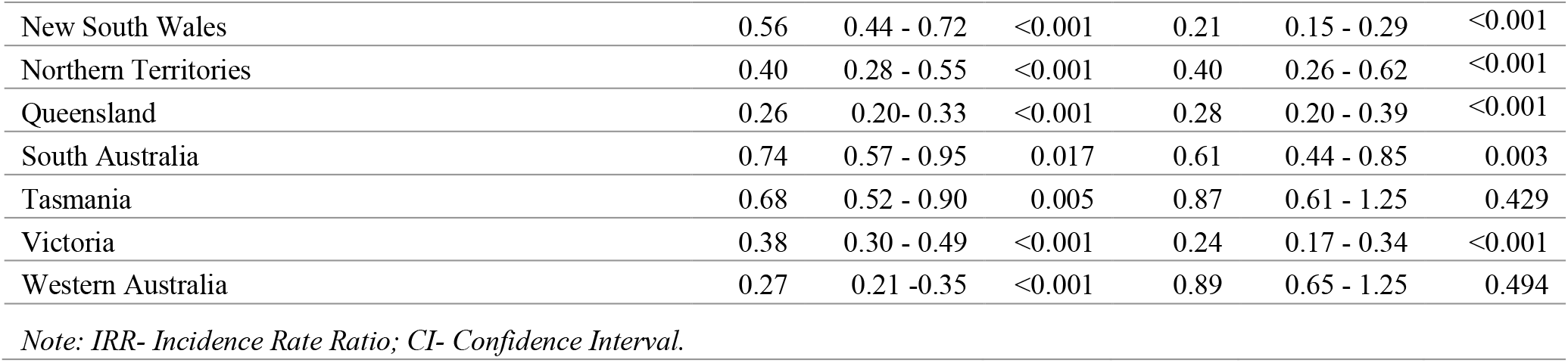
Factors associated with risk of mental health conditions and assault-injury among educators.

| Injury | Mental Health Conditions |  |  | Assault-injury |  |  |
| --- | --- | --- | --- | --- | --- | --- |
|  | IRR | 95% CI | p-value | IRR | 95% CI | p-value |
| **Educator subgroups** (reference: Secondary Education) |  |  |  |  |  |  |
| Childcare and Early Learning | 0.39 | 0.33 - 0.46 | <0.001 | 0.77 | 0.62 - 0.96 | 0.020 |
| Primary Education | 0.63 | 0.55 - 0.71 | <0.001 | 0.70 | 0.58 - 0.85 | <0.001 |
| Principals | 0.67 | 0.56 - 0.80 | <0.001 | 1.19 | 0.91 - 1.55 | 0.189 |
| Tertiary Education | 0.14 | 0.11 - 0.17 | <0.001 | 0.07 | 0.03 - 0.13 | <0.001 |
| Vocational Education | 0.50 | 0.42 - 0.60 | <0.001 | 0.20 | 0.12 - 0.30 | <0.001 |
| Special Education | 1.17 | 0.98 - 1.40 | 0.082 | 8.46 | 6.93 - 10.31 | <0.001 |
| Education Aides | 0.71 | 0.61 - 0.83 | <0.001 | 3.68 | 3.05 - 4.43 | <0.001 |
| **Sex** (reference: Female) |  |  |  |  |  |  |
| Male | 0.91 | 0.83 - 1.01 | 0.071 | 0.86 | 0.75 - 0.99 | 0.028 |
| **Age group** (reference: 18-24 years) |  |  |  |  |  |  |
| 25-29 years | 2.03 | 1.57 - 2.64 | <0.001 | 1.15 | 0.88 - 1.52 | 0.306 |
| 30-34 years | 2.42 | 1.88 - 3.13 | <0.001 | 1.20 | 0.92 - 1.58 | 0.181 |
| 35-39 years | 3.02 | 2.36 - 3.88 | <0.001 | 1.13 | 0.86 - 1.48 | 0.378 |
| 40-44 years | 3.77 | 2.96 - 4.82 | <0.001 | 1.32 | 1.01 - 1.72 | 0.038 |
| 45-49 years | 4.93 | 3.88 - 6.29 | <0.001 | 1.67 | 1.29 - 2.18 | <0.001 |
| 50-54 years | 5.30 | 4.18 - 6.74 | <0.001 | 1.51 | 1.17 - 1.97 | 0.002 |
| 55+ years | 4.60 | 3.64 - 5.84 | <0.001 | 1.31 | 1.02 - 1.70 | 0.035 |
| **Jurisdiction** (reference: Comcare) |  |  |  |  |  |  |
| New South Wales | 0.56 | 0.44 - 0.72 | <0.001 | 0.21 | 0.15 - 0.29 | <0.001 |
| Northern Territories | 0.40 | 0.28 - 0.55 | <0.001 | 0.40 | 0.26 - 0.62 | <0.001 |
| Queensland | 0.26 | 0.20 - 0.33 | <0.001 | 0.28 | 0.20 - 0.39 | <0.001 |
| South Australia | 0.74 | 0.57 - 0.95 | 0.017 | 0.61 | 0.44 - 0.85 | 0.003 |
| Tasmania | 0.68 | 0.52 - 0.90 | 0.005 | 0.87 | 0.61 - 1.25 | 0.429 |
| Victoria | 0.38 | 0.30 - 0.49 | <0.001 | 0.24 | 0.17 - 0.34 | <0.001 |
| Western Australia | 0.27 | 0.21 - 0.35 | <0.001 | 0.89 | 0.65 - 1.25 | 0.494 |

*Note: IRR- Incidence Rate Ratio; CI- Confidence Interval.*

### Duration of time loss

Educators had shorter durations of time loss overall compared to non-educators (Hazard Ratio [HR]: 1.17; 95% CI: 1.10-1.24). Overall, duration of time loss within educators did not differ significantly from their reference categories for age group, injury type, mechanism of injury, remoteness, and socioeconomic advantage. Within educator subgroups, education aides had the longest disability duration (HR: 0.70; 95% CI: 0.56-0.87). Otherwise, no other educator group differed significantly from the reference category (secondary education). Shortest duration of time loss was in Queensland (HR: 4.35; 95% CI: 2.51-7.54). There were no detectable differences based on remoteness and socioeconomic status. (Table 5).

**Table 5:** Factors associated with educators’ duration of time loss in weeks, results of Cox regression analyses with multiple imputation for SES and remoteness.

| ***Educators compared to non-educators*** | **HR\*** | **95% CI** | **p-value** |
| --- | --- | --- | --- |
| **Occupation** (reference: Non-Educators) |  |  |  |
| Educators | 1.17 | 1.10 - 1.24 | <0.001 |
| **Within Educators Analysis** |  |  |  |
| **Educator Subgroups** (reference: Secondary Education) |  |  |  |
| Childcare and Early Learning | 0.87 | 0.72 - 1.06 | 0.178 |
| Primary Education | 0.94 | 0.77 - 1.14 | 0.510 |
| Principals | 0.95 | 0.64 - 1.40 | 0.781 |
| Tertiary Education | 0.79 | 0.53 - 1.19 | 0.264 |
| Vocational Education | 0.90 | 0.68 - 1.19 | 0.456 |
| Special Education | 0.90 | 0.67 - 1.21 | 0.483 |
| Education Aides | 0.70 | 0.56 - 0.87 | 0.002 |
| **Sex** (reference: Female) |  |  |  |
| Male | 0.99 | 0.84 - 1.16 | 0.908 |
| **Age Group** (reference: 18-24 years) |  |  |  |
| 25-29 years | 1.07 | 0.63 - 1.80 | 0.813 |
| 30-34 years | 1.27 | 0.77 - 2.09 | 0.345 |
| 35-39 years | 0.96 | 0.58 - 1.57 | 0.867 |
| 40-44 years | 1.02 | 0.64 - 1.61 | 0.946 |
| 45-49 years | 1.01 | 0.63 - 1.59 | 0.980 |
| 50-54 years | 1.00 | 0.63 - 1.57 | 0.990 |
| 55+ years | 1.35 | 0.86 - 2.13 | 0.198 |
| **Injury** (reference: Fractures) |  |  |  |
| Upper Body Musculoskeletal | 0.94 | 0.69 - 1.28 | 0.700 |
| Lower Body Musculoskeletal | 1.10 | 0.78 - 1.57 | 0.580 |
| Other Musculoskeletal | 0.97 | 0.65 - 1.46 | 0.900 |
| Mental Health Conditions | 0.86 | 0.64 - 1.17 | 0.350 |
| Neurological | 1.07 | 0.65 - 1.77 | 0.790 |
| Other Diseases | 0.65 | 0.42 - 1.00 | 0.050 |
| Other Traumatic | 0.84 | 0.59 - 1.22 | 0.360 |
| Other/Unidentified | 5.12 | 0.68 - 38.83 | 0.110 |
| **Mechanism of injury** (reference: Not Assault) |  |  |  |
| Assault | 1.28 | 0.93 - 1.76 | 0.132 |
| **Jurisdiction** (reference: Comcare) |  |  |  |
| New South Wales | 1.57 | 0.99 - 2.50 | 0.054 |
| Northern Territories | 2.62 | 0.96 - 7.13 | 0.060 |
| Queensland | 4.35 | 2.51 - 7.54 | <0.001 |
| South Australia | 2.23 | 1.36 - 3.67 | 0.001 |
| Tasmania | 1.36 | 0.79 - 2.36 | 0.269 |
| Victoria | 1.87 | 1.18 - 2.95 | 0.007 |
| Western Australia | 3.85 | 2.34 - 6.34 | <0.001 |
| **Remoteness** (reference: Major Cities of Australia) |  |  |  |
| Inner Regional Australia | 1.06 | 0.90 - 1.25 | 0.486 |
| Outer Regional Australia | 0.90 | 0.69 - 1.19 | 0.474 |
| Remote Australia | 0.87 | 0.48 - 1.58 | 0.644 |
| Very Remote Australia | 0.84 | 0.35 - 1.98 | 0.685 |
| **Socioeconomic Status** (reference: Disadvantaged) |  |  |  |
| Middle 3 Quintiles | 1.01 | 0.84 - 1.21 | 0.933 |
| Most Advantaged | 0.99 | 0.80 - 1.23 | 0.927 |

Note: HR - Hazard Ratio; CI- Confidence Interval; SES - Socioeconomic Status.
\*HR<1 indicates longer disability duration, HR> 1 indicates shorter duration of time loss.

## Discussion

### Rate of injury claims

Th findings suggest that educators are at lower risk of injury resulting in a workers’ compensation claim than non-educators. This is in contrast to survey data that captures all self-reported work-related conditions, regardless of whether they were compensated. In 2017/18, an estimated 53 per 1000 workers in the education and training industry experienced work-related injury or illness, the third highest rate among Australian industries [39]. However, claims data may systematically underestimate occupational injuries and illnesses in educators relative to other occupations due to the absence-discouraging culture of their work environment, leadership attitude, inclination to work when advised against it, and utilizing fixed school holidays to de-stress and recuperate [40,41]. Taking time off work also entails lesson planning for substitutes, which could also discourage educators from lodging a workers’ compensation claim.

Despite the likely underreporting, educators had higher rates of compensable mental health conditions and injuries arising from assault, in concordance with the existing evidence [8,14–23]. Also consistent with previous studies, musculoskeletal injuries accounted for the majority of claims [2–12], though educators had a lower rate of these claims compared to non-educators. It is notable that while education is a female dominated sector, this study found no significant difference between male and female claim rates.

Consistent with the current evidence, educators in secondary education had the highest injury claim rates [7,8,11,19]. Special educators had the highest rate of mental health injury claims; while not significantly higher than the reference group, secondary educators, all other educator subgroups were significantly lower, which suggests educators in special education have an especially high risk of mental health conditions. This may be due to the unique challenges of their work including special needs of their students, limited resources, rigid curriculum, and insufficient support [42,43]. Additionally, a review of the Australian special educators’ training pathways found inconsistencies that may have led to gaps in some educators’ management of challenging classroom behavior [42]. These challenges likely contribute to educators’ stress, discouragement, frustration, poor coping behaviors, and increased risk of burnout [43–45].

The finding that educators, particularly those in special education and education aides, were at elevated risk of compensable injuries resulting from assault is consistent with previous research [21,24–26,46–49]. Special educators and education aides often work with students who have physical and behavioral needs, adding more responsibility to their role than typical educators, like managing the classroom and student safety, providing individualized support, and closely working with the students throughout the school day [21]. Violence is evidenced to have an immense impact on educators’ quality of life and overall wellbeing, including increased risk of cardiovascular disease and sleep disorders, decreased sense of safety, and poor mental and emotional wellbeing [16,26,50,51].

Educators may refrain from reporting incidents in some situations because they may assume students are acting out due to distress and their actions are unintended [26]. The possibility that educators underreport occupational injuries and illnesses makes their elevated rate of claims for mental health and assault all the more striking and suggests they are severe underestimates.

### Disability duration

The shortened disability duration among educators could reflect less severe injuries and faster recovery within this group. Alternatively, educators may have truncated periods on compensation and return to work earlier in the recovery process. Some educators asserted that they would prefer taking short breaks to rest in order to avoid extended leave periods [41]. Such recovery times would not be captured by compensation databases.

Educators may also face institutional and systematic challenges to taking compensated time off work after injury. Those who are frequently absent may be stigmatized, ostracized, and distrusted by their colleagues and administration [41]. Conversely, evidence suggests decreased absenteeism is not an appropriate measure of good health, as educators were three to four times more likely to be present at work while sick than managers [52]. Thus, disability duration may not be the best measure to assess the impact of educators’ work-related injuries and illness.

The shortest disability durations were in Queensland and Western Australia, which may be due to the alleged high prevalence of injuries of an acute nature, like those relating to assault [28,53]. It may also be attributable to differences of compensation systems and eligibility among jurisdictions, as a lower threshold leads to shorter average disability durations [54,55].

SES and location are used to denote resource availability and service accessibility, and have been linked to disability duration in previous research [34]. While most educators’ injury claims originated from major cities and middle SES, this study found no association with disability duration.

### Strengths and limitations

Study strengths include the use of population-level claims data derived from 8 of the 11 national compensation schemes’ data, covering 90% of the Australian workforce [34], facilitating effective comparison of claims and injury rates among occupation groups.

This study is limited by using the NDS dataset, which is collected for administrative rather than research purposes. This may introduce systematic bias which can distort injury risk between occupations. The limited dataset does not allow exploration of potential psychosocial confounding variables. Compensation data underestimates the prevalence of work-related injuries and diseases and the actual time off work, which may also systematically differ between educators and non-educators.

The COVID-19 pandemic in 2020 has caused considerable disruptions requiring the adaptation of a hybrid education system in which educators are working in-person and online. The increased prevalence of remote education can lead to additional injury risks that were not able to be covered in this paper, due to the timeframe of data, however, it might be worthwhile repeating the study in the future to determine the differences.

## Conclusion

While educators had lower rates of injury claims and compensated disability duration, the inconsistency with work injury surveys suggests underreporting of injuries to the compensation system. Regardless, educators had higher rates of injury claims for mental health conditions and assault. This study provided insight into injury prevalence and type, identified risk categories, and highlighted the vulnerability of educator subgroups, in particular those in special education and education aides. Educators’ risk of assault and mental health conditions is concerning, suggesting the need for preventive interventions. Future research could explore factors associated with educators’ injuries and how they can be effectively targeted.

## Data Availability

All data in the present work are potentially identifiable workers' compensation claim records. Our data sharing agreement with Safe Work Australia and the compensation systems throughout Australia forbid sharing.

### Appendix 1

| ANZSO educators' occupation codes and their regrouping in this study |  |  |
| --- | --- | --- |
| Code | Occupation | Group in this study |
| 2411 | Early Childhood Teachers | Childcare and Early Learning |
| 4211 | Child Careers |  |
| 2412 | Primary School Teachers | Primary Education |
| 2413 | Middle School Teachers | Secondary Education |
| 2414 | Secondary School Teachers |  |
| 1343 | School Principal | Principals |
| 2421 | University Lecturers and Tutors | Tertiary Education |
| 2422 | Vocational Education Teachers | Vocational Education |
| 2415 | Special Education Teachers | Special Education |

## Notes

Declarations of interest The Authors declare that there is no conflict of interest.

### Competing Interest Statement

The authors have declared no competing interest.

### Funding Statement

This study did not receive any funding

### Author Declarations

The Monash University Human Research Ethics Committee (MUHREC) of Monash University gave ethical approval for this work (CF14/2995 2014001663).

